# The Impact of Sleep on Breast Cancer-Specific Mortality: A Mendelian Randomisation Study

**DOI:** 10.1101/2023.06.07.23291014

**Authors:** Bryony L Hayes, Leanne Fleming, Osama Mahmoud, Richard M Martin, Deborah A Lawlor, Timothy Robinson, Rebecca C Richmond

**Affiliations:** Medical Research Council Integrative Epidemiology Unit, University of Bristol, Bristol, UK; Population Health Sciences, Bristol Medical School, University of Bristol, Bristol, UK; University of Strathclyde, George Street, Glasgow, UK; Department of Mathematical Sciences, University of Essex, Colchester, UK; Department of Applied Statistics, Helwan University, Helwan, Egypt; NIHR Bristol Biomedical Research Centre, Bristol, UK; Bristol Cancer Institute, University Hospitals Bristol and Weston NHS Foundation Trust, Bristol, UK

## Abstract

The relationship between sleep traits and survival in breast cancer is uncertain and complex. There are multiple biological, psychological and treatment-related factors that could link sleep and cancer outcomes. Previous studies could be biased due to methodological limitations such as reverse causation and confounding. Here, we used two-sample mendelian randomisation (MR) to investigate the causal relationship between sleep and breast cancer mortality.

Publicly available genetic summary data from females of European ancestry from UK Biobank and 23andme and the Breast Cancer Association Consortium were used to generate instrumental variables for sleep traits (chronotype, insomnia symptoms, sleep duration, napping, daytime-sleepiness, and ease of getting up (N= 446,118-1,409,137)) and breast cancer outcomes (15 years post-diagnosis, stratified by tumour subtype and treatment (N=91,686 and Ndeaths=7,531 over a median follow-up of 8.1 years)). Sensitivity analyses were used to assess the robustness of analyses to MR assumptions.

Initial results found some evidence for a per category increase in daytime-sleepiness reducing overall breast cancer mortality (HR=0.34, 95% CI=0.14, 0.80), and for insomnia symptoms reducing odds of mortality in oestrogen receptor positive breast cancers not receiving chemotherapy (HR=0.18, 95% CI=0.05, 0.68) and in patients receiving aromatase inhibitors (HR=0.23, 95% CI=0.07, 0.78). Importantly, these relationships were not robust following sensitivity analyses meaning we could not demonstrate any causal relationships.

This study did not provide evidence that sleep traits have a causal role in breast cancer mortality. Further work characterising disruption to normal sleep behaviours and its effects on tumour biology, treatment compliance and quality of life are needed.

## INTRODUCTION

There were an estimated 2.3 million new cases of breast cancer worldwide in 2020 and 685,000 breast cancer specific deaths, making it the leading cancer and cause of cancer death in females[1]. Breast cancer is a heterogeneous disease and prognosis varies according to a patient’s age and stage at diagnosis [2,3], histological subtype [4,5], lifestyle factors [6,7] and treatment received. Severely disrupted sleep is a common complaint among breast cancer patients, with ∼20–70% of breast cancer patients reporting insomnia, defined as chronic difficulty (3 months or more) with sleep initiation or maintenance, resulting in daytime functional impairment [8,9]. Insomnia following a breast cancer diagnosis is often long-term and has a negative impact on both the quality of life of breast cancer survivors as well as adherence to treatment regimen [10–15]. It is also often associated with a symptom cluster of depression, anxiety and fatigue [16].

It is likely that both psychological aspects of breast cancer diagnosis (e.g. anxiety [17]) as well as the hormonal effects of treatment (e.g. side-effects from prescribed medications such as selective oestrogen receptor modifiers (SERMs) and aromatase inhibitors (AIs)) [18]) contribute to the sleep disruption experienced by breast cancer patients. Furthermore, trials of both tamoxifen (a SERM) and exemestane (an AI), found that sleep disruption can continue up to 24 months post-diagnosis, with some evidence for continued disruption up to 60 months [19]. This may indirectly impact breast cancer survival, for example through low adherence to treatment and higher rates of relapse (early discontinuation of treatment plan results in a 20-50% increased risk of mortality [14,20]), or there may be a direct mechanism by which sleep influences survival chances.

Nevertheless, the relationship between sleep traits and breast cancer survival is unclear. A previous observational study of 3682 women with stage I and II breast cancer found that prolonged (rather than disrupted) sleep had a detrimental effect on survival, where women sleeping ≥9 hours per night had a higher risk of breast cancer mortality (hazard ratio (HR)=1.46, 95% confidence interval (CI)=1.02,2.07) compared to women sleeping 8 hours per night, and no evidence for an association between short sleep and mortality[21]. In support of this, a recent mechanistic study conducted in a patient group (N=30), with further *in-vivo* work in mouse cancer models, found an increase in circulating tumour cells (CTCs) that led to subsequent metastases and upregulation of mitotic genes that occurred almost exclusively during sleep [22], suggesting that longer sleep may increase risk of metastases.

As well as confirming the adverse effect of longer sleep duration, a recent systematic review of cancer outcomes in breast cancer survivors found some evidence to suggest that measures of better sleep quality are associated with lower risk of all-cause mortality (HR range= 0.29-0.97) and shorter nightly fasting duration, as a proxy for circadian timing, was associated with higher risk of all breast cancer outcomes (HR range= 1.21-1.36) [23].

Due to the limited number of studies to date, and potential for residual confounding explaining associations in observational studies, further evidence of the potential effect of sleep on breast cancer survival is warranted. Mendelian randomisation (MR) is an analytical approach that uses germline genetic variants as instrumental variables (IVs) for potentially modifiable risk factors to assess their causal effect on an outcome [24,25]. Provided a specific set of assumptions are met, MR incurs less bias from factors such as non-genetic confounding and reverse causation compared to conventional observational analyses [24–26]. This approach has been traditionally used to evaluate causal effects of exposures on disease incidence. We have previously used MR to provide evidence of a protective effect of morning-preference chronotype (odds ratio (OR)=0.88; 95% CI= 0.82, 0.93 per category increase) and an adverse effect of long sleep duration (OR= 1.19; 95% CI= 1.02, 1.39 per hour increase) on breast cancer risk [27].

MR can be also used to study factors that potentially influence disease prognosis. A recent MR study assessed the effect of 9 established risk factors for breast cancer occurrence on breast cancer survival as an outcome, finding robust evidence that type 2 diabetes reduced survival but other aetiological risk factors did not influence survival [28]. The effect of sleep traits on breast cancer survival was not included in that study and we are not aware of any other MR studies exploring their effects on breast cancer survival.

The aim of this study was to explore the potential effect of selected sleep traits capturing sleep duration, sleep quality/disturbance (insomnia symptoms, daytime sleepiness) and circadian timing (chronotype, napping during the day, and ease of getting up in the morning) on breast cancer specific mortality (over a median follow up of 8.1 years, up to 15-years post-diagnosis) using two-sample MR and leveraging the largest genome-wide association study (GWAS) on breast cancer survival recently released by the Breast Cancer Association Consortium (N= 91,686) [29], with further exploratory analyses utilising data stratified by breast cancer subtype and treatment.

## METHODS

### Study Design

We performed two-sample MR analyses in which the associations of germline genetic variants with the exposure (sleep trait) and the outcome (breast cancer mortality) were estimated in two independent (i.e. non-overlapping) samples. The sleep traits explored in this study were: sleep duration, insomnia symptoms, daytime sleepiness, and morning preference chronotype, napping during the day, and ease of getting up in the morning). The outcome in this study was breast cancer specific mortality (has died/has not died) up to 15-years post-diagnosis.

### Study Samples

#### Sleep Traits in UK Biobank

Genetic instruments (single nucleotide polymorphisms, SNPs) for morning preference, sleep duration, napping during the day, daytime sleepiness and getting up in the morning were selected from published genome wide association studies (GWAS) that included adult women and men from the UK Biobank [30–33].

Between 2006 and 2010, UKB recruited >500,000 participants (55% women) between 40 and 70 years of age out of the 9.2 million invited to participate (5.5% response rate)[34]. Details on recruitment and informed consent have been published previously[35]. At a baseline visit to assessment centres, participants completed a touchscreen questionnaire, which included questions about their sleep behaviours (details of these questions are available in **Supplementary Methods A**).

To obtain estimates for the SNP-exposure association (sample 1), we conducted our own GWAS for each of these sleep traits in UK Biobank (UKB) in women only. This was done using a linear mixed model (LMM) method to account for relatedness and population stratification, as implemented in BOLT-LMM (v2.3) [36]. More details of this approach can be found in **Supplementary Methods B**.

From our GWAS results, we extracted the female-specific effect estimates for the genetic variants identified in the relevant published sleep GWAS (**supplementary data Table A**). These effect estimates were used regardless of whether the female-specific association reached genome-wide significance (GWS) (p<5x10-8) in our UKB analysis. This approach was taken for all sleep traits, with the exception of insomnia symptoms.

For insomnia, female-specific SNP effects for self-reported insomnia symptoms have been previously reported in a published meta-GWAS conducted in UKB and 23andMe[37] (details of 23andMe and the reporting of insomnia symptoms in that study are available in **Supplementary Methods C**). In contrast to the other sleep traits, the female-specific effect estimates for insomnia symptoms had accompanying p-values which reached GWS (p<5x10-8).

#### Breast Cancer Mortality in BCAC

All breast cancer mortality data were generated from women with breast cancer who were participants in the Breast Cancer Association Consortium (BCAC)[29]. BCAC is a GWAS consortium comprising 70 breast cancer studies with a total of 122,977 cases and 105,974 controls. The GWAS of breast-cancer specific deaths involved 91,686 women with breast cancer, of whom 7,531 died of their breast cancer over a median follow-up of 8.1 years [29]. Detailed information for the included studies has been provided elsewhere[29].

All studies for sleep and breast cancer data were approved by the relevant ethics committees and informed consent was obtained from all participants. All participants included in the sleep and breast cancer GWAS were of European ancestry.

### Constructing Genetic Instruments

For chronotype, 351 SNPs were identified in the largest published GWAS (N = 697,828)[30], of which 341 were present in the imputed female UKB-only GWAS that was conducted for this paper. For sleep duration (N = 446, 118)[31] 91 SNPs had previously been identified, all of which were present in our female-only GWAS. For napping (N = 452,633)[32], 123 SNPs had previously been identified, of which 94 were present in our female-only GWAS. For daytime sleepiness (N = 452,071)[33], 42 SNPs had previously been identified, of which 32 were present in our female-only GWAS. For ease of getting up in the morning (N = 461,658)[33], 76 SNPs had previously been identified, all of which were present in our female-only GWAS. For insomnia symptoms, there were 953 SNPs identified in the published female-only meta-GWAS of UKB and 23andme (N = 1,409,137)[37].

These sleep trait instruments were then subjected to LD-clumping to obtain independent SNP-associations (using a threshold of R2 = 0.001). **Supplementary Data Table A** provides details of the SNPs used to instrument each of the sleep traits in this study.

### Statistical Analyses

The TwoSampleMR R package (version 0.5.6[26], R version 4.0.4.) was first used to combine and harmonise summary genetic data for the sleep exposure traits with the breast cancer mortality GWAS. During harmonisation, palindromic SNPs can be aligned when the minor allele frequency is <0.3, and are otherwise excluded. For sleep SNPs not present in the breast cancer outcome GWAS, proxy SNPs were identified (linkage disequilibrium r^2^ > 0.8).

We used inverse variance weighted (IVW) MR, whereby an aggregate estimate of the causal effect is obtained from the slope of the weighted instrumental variable (IV)-mean exposure regressed against the IV-mean outcome associations, with an intercept of zero.

#### Sensitivity Analyses and Limiting Assumption Violation

To ensure appropriate causal inference and validity in an MR study, three key assumptions must be met: i) genetic IVs must be robustly associated with the exposure of interest, within the target population (relevance assumption); ii) there must be no confounding between genetic IVs and the outcome being tested (independence assumption); and iii) genetic IVs may only affect the outcome via the exposure of interest (exclusion restriction assumption)[24].

F-statistics were used to measure the strength of the relationship between genetic IVs and corresponding sleep trait exposures to satisfy the relevance assumption[38,39], and r^2^ was used to explain the proportion of variance in sleep traits explained by our instruments. To mitigate against confounding by population stratification, the most likely source of confounding in MR studies, we restricted our analysis to genetic data derived for Europeans only. Horizontal pleiotropy – where the genetic instruments associate with the outcome (here death) independently of the exposure of interest (sleep traits) – is the most common cause of violation of the exclusion restriction criteria. We assessed this using a series of sensitivity analyses. First, as heterogeneity in the causal effect estimated between SNPs (between-SNP heterogeneity) is an indicator of some genetic instruments being pleiotropic, we used Cochran’s Q and I^2^ statistics to explore this [25,40,41]. To identify SNPs with the largest contribution towards heterogeneity, radial-MR was conducted using modified second order weights and an alpha level of 0.05 divided by the number of SNPs being used to instrument the exposure [42]. Following radial-MR, analyses were repeated with any flagged outlier SNPs removed to assess the impact of their potential bias.

MR was conducted using MR-Egger[43], weighted median[44] and weighted mode[45] to assess bias from unbalanced pleiotropy. To assess expected relative bias of the MR-Egger causal estimate, weighted and unweighted I^2^_GX_ statistics were calculated [46], and to extrapolate bias-adjusted inference, SIMEX corrections were subsequently conducted if I^2^_GX_ were sufficiently low (less than 90%) and therefore indicating potential violation of the ‘NO Measurement Error’ (NOME) assumption [47].

Additional considerations must also be made when performing two-sample MR analysis of disease progression studies. For two-sample MR, it is assumed that the two samples included are independent of each other. Since sample overlap between UKB and BCAC is negligible, analyses between these samples should not violate this assumption. It is also assumed that the two samples are drawn from the same population. In support of this, we used female- and European-only GWAS data for the exposures and outcome.

Within the context of disease progression studies, MR may be affected by index event bias (a form of collider bias) which can occur if the exposure of interest (e.g. sleep trait) associates with both disease occurrence and prognosis in those with disease (i.e. both breast cancer incidence and mortality), and there is one or more other factors that associate with disease occurrence and prognosis (**Figure 1**)[48]. For example, previous MR studies suggest type 2 diabetes influences breast cancer occurrence and survival, and sleep traits have been previously found to be causally related to breast cancer incidence [27], and type 2 diabetes traits [49]. Hence when we use MR to assess survival only in women with breast cancer, a spurious association may be generated via type 2 diabetes (**Figure 1**).

**Figure 1.**
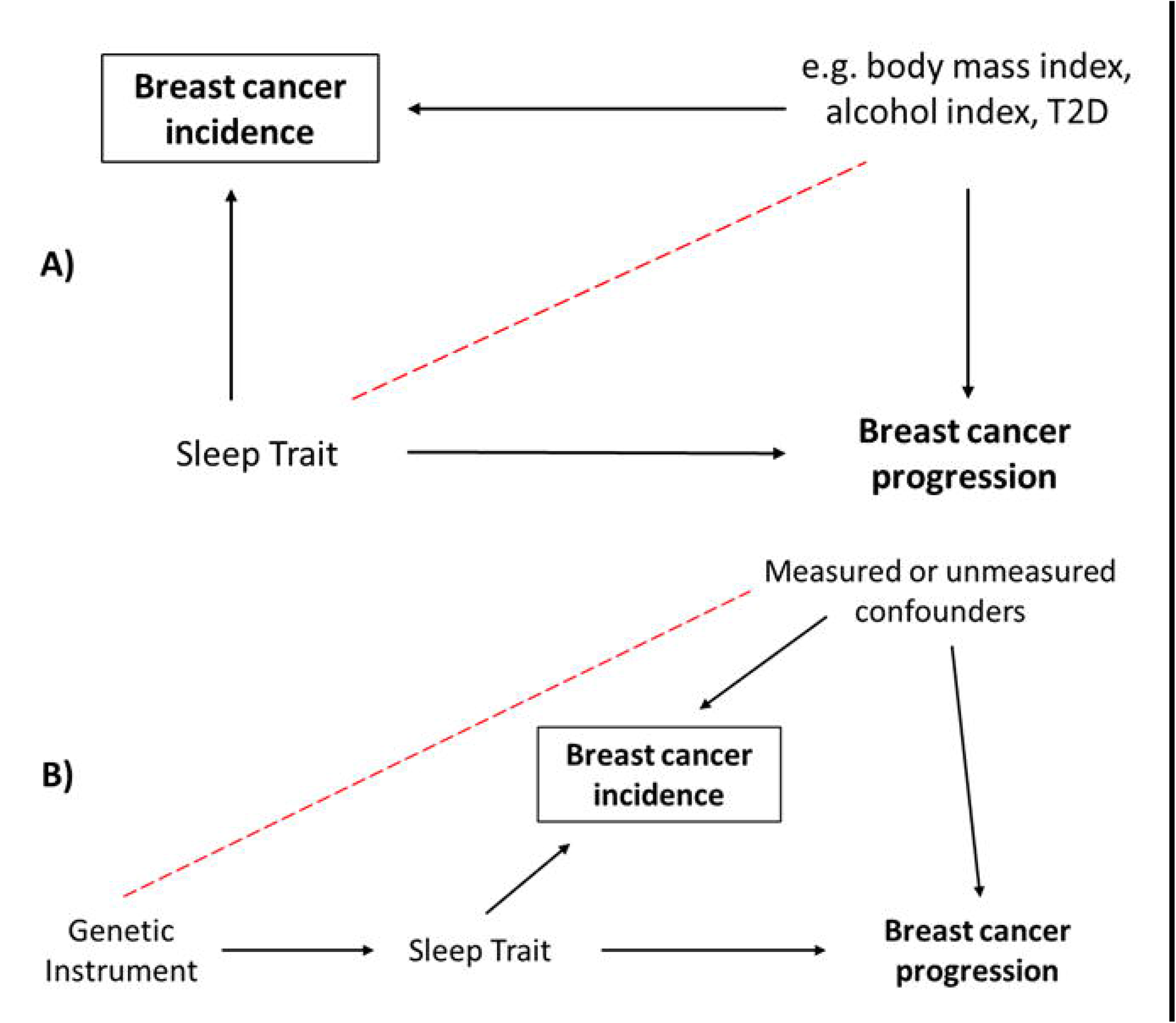
A) Direct acyclic graph demonstrating the introduction of collider bias. B) Collider bias in Mendelian randomisation analyses, where the exposure is proxied by a genetic instrument.

We first performed two-sample MR analysis of the six sleep traits in relation to breast cancer incidence to establish which traits may be influenced by collider bias in the MR analysis. Any sleep traits found to have an effect on breast cancer incidence were then investigated to determine the influence of collider bias. This was done using the same SNP-sleep trait associations (Supplementary Data Table A) as in the main analysis (sample 1) and SNP-breast cancer incidence associations (sample 2) from a BCAC incidence GWAS comprised of 67 studies (N cases = 133,384; N controls = 113,789) [50].

We then used the Slope-Hunter method (V1.1.0 [51]), which estimates the effect of our genetic instruments on progression with adjustment for any effect of the instruments on incidence [52]. [52]This was applied using both the GWAS for “all patients” 15-year breast cancer mortality and the GWAS of breast cancer incidence within BCAC [50] to partition SNPs into 1) SNPs only affecting incidence and 2) SNPs affecting both incidence and progression. A correction factor was estimated from the slope of the regression line of disease progression associations on disease incidence associations using the SNPs only affecting incidence, and this correction factor was then applied to all SNPs in the breast cancer mortality GWAS. We then re-ran the two-sample MR analysis for those sleep traits found to be causally related to breast cancer incidence using this corrected mortality GWAS.

#### Supplementary Analysis

While our main outcome was overall breast-cancer specific mortality at 15 years, further exploratory analyses was conducted using mortality GWAS for breast cancer subgroups stratified by tumour type and treatment (**Table 1**). The scientific justification to support the stratification of the mortality results by each breast cancer subgroup has been described in detail elsewhere[29]. With respect to our analysis, we were interested in investigating potential effect modification by tumour- or treatment-type, given the disruptive effect certain therapies (e.g., aromatase inhibitors) have on sleep. We assessed this using I^2^ (%) from meta-analysis of causal estimates to determine between-group heterogeneity [53], in which one meta-analysis comprised of patients stratified by both tumour-type and treatment, and the other of patients stratified by treatment-only.

**Table 1.** Breast cancer subgroups, sample sizes and number of deaths. ER = Oestrogen receptor, PR = Progesterone receptor, HER2 = Human epidermal growth factor receptor 2.

| Group Description | N | N Deaths |
| --- | --- | --- |
| All patients (15 years) | 91686 | 7531 |
| **Tumour-type and treatment** |  |  |
| ER+ tumours (endocrine therapy) | 30137 | 2298 |
| ER- tumours (chemotherapy) | 6852 | 907 |
| ER+/PR+ HER2- tumours (Total) | 35550 | 2141 |
| ER+/PR+ HER2- tumours (chemotherapy) | 10394 | 889 |
| ER+/PR+ HER2- tumours (no chemotherapy) | 10275 | 457 |
| ER+/PR+ HER2+ tumours (all therapies) | 5567 | 520 |
| ER-/PR- HER2+ tumours (all therapies) | 2298 | 314 |
| ER-/PR- HER2- tumours (all therapies) | 5631 | 677 |
| **Treatment-only** |  |  |
| All patients given tamoxifen | 17327 | 1777 |
| All patients given aromatase inhibitor | 9041 | 495 |
| All patients given CMF-like Chemo | 3044 | 381 |
| All patients given taxanes | 6007 | 462 |
| All patients given anthracyclines | 12475 | 1292 |

All results were subjected to Bonferroni correction to account for type 1 error due to multiple testing. The corrected P-value threshold was calculated from original P-value threshold divided by number of tests undertaken (main analyses: (0.05/6) = 0.0083; supplementary analyses: (0.05/14x6) = 0.0006).

## RESULTS

### Main Analysis

We found little evidence for an effect of any of the measures of sleep on overall breast cancer-specific mortality during 15-years’ follow-up: HR= 1.05, 95% CI= 0.91, 1.20 per category increase in chronotype (from definitely an ‘evening person’ to definitely a ‘morning person’); HR= 0.95, 95% CI= 0.70, 1.29 in participants experiencing insomnia symptoms (compared to no symptoms); HR= 1.08, 95% CI= 0.82, 1.41 per hour increase in sleep duration; HR= 0.94, 95% CI= 0.60, 1.48 per category increase in napping (from never/rarely to usually); and HR= 1.03, 95% CI= 0.69, 1.53 per category increase in ease of getting up (from not at all easy to very easy) (**Figure 2**).

**Figure 2.**
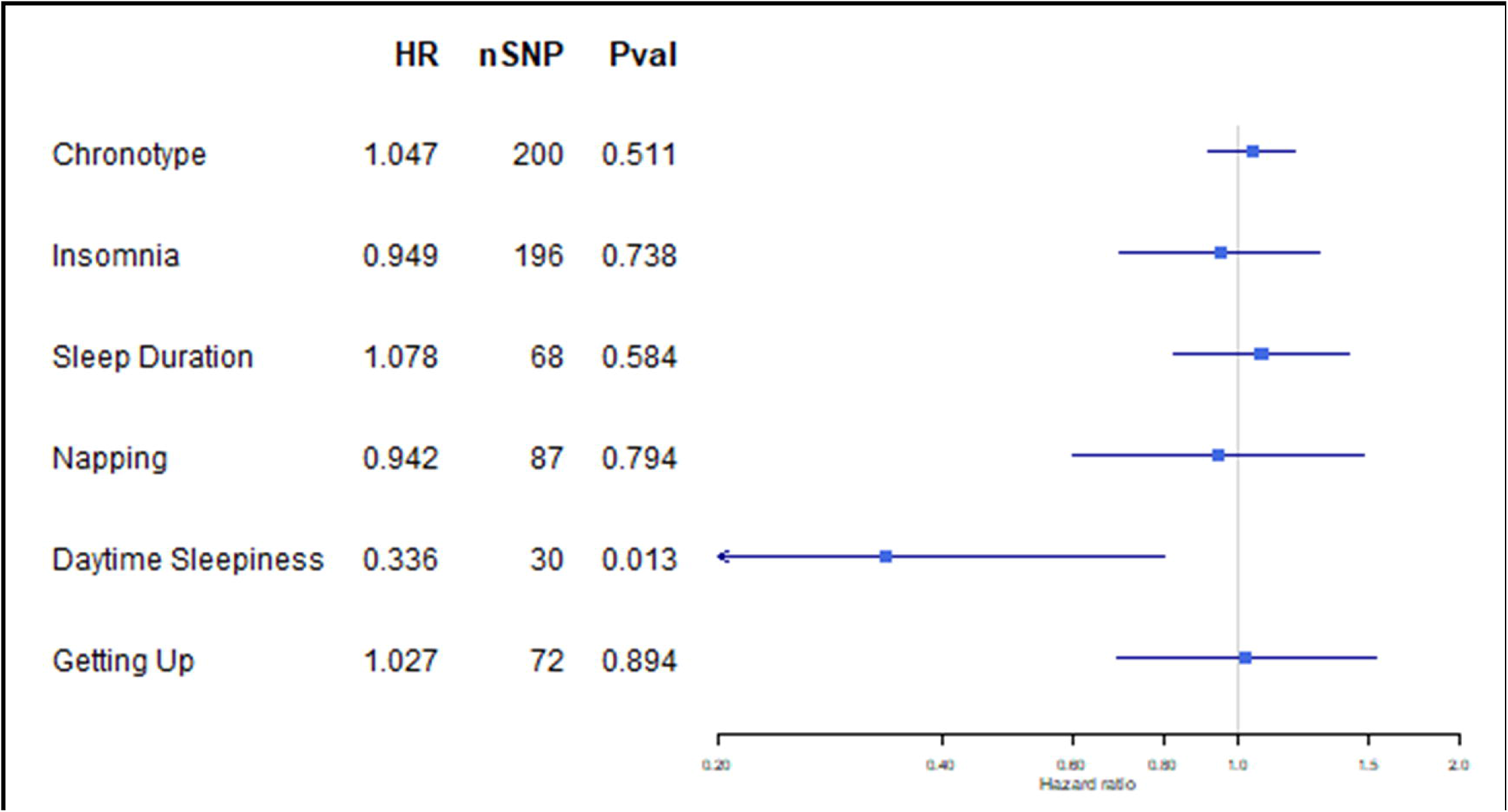
Two-sample MR results for sleep effect on breast cancer mortality in all patients at a 15-year timepoint.

We found suggestive evidence that a per category increase in daytime sleepiness (from never/rarely to often) reduced the odds of breast cancer specific mortality (HR= 0.34, 95% CI= 0.14, 0.80, p=0.013), although this result was not robust to Bonferroni correction to adjust for multiple testing (Pval threshold = 0.0083) (**Figure 2**).

### Sensitivity Analysis

Genetic variants contributing to the sleep trait instruments had a combined F-statistic of 22.83-65.25, indicating reasonable instrument strength [29,40]. R^2^ values were calculated for all sleep traits and suggested that the instruments explained 0.35-1.97% of the variance of the exposures. The highest between-SNP heterogeneity among sleep trait instruments was I^2^=24% (Qstat = 93.40; Qpval = 0.037) for ease of getting up in the morning, indicating low to moderate heterogeneity and therefore low likelihood of pleiotropy. In our main analyses, no outlier SNPs were removed during radial MR. Comparisons between IVW, MR-Egger and weighted median were largely consistent for our main analyses, although MR-Egger estimates were particularly imprecise (**supplementary tables B to G**).

Weighted and unweighted I2GX values [36] were also calculated for each of the sleep traits on mortality outcomes and found to be between 96-98%, indicating no NOME violation in our MR-Egger analyses. Therefore, SIMEX corrections [37] were not conducted for these analyses. Results of these sensitivity analyses have been summarised in **Table 2**, and complete results can be found in **supplementary table H**.

**Table 2.**
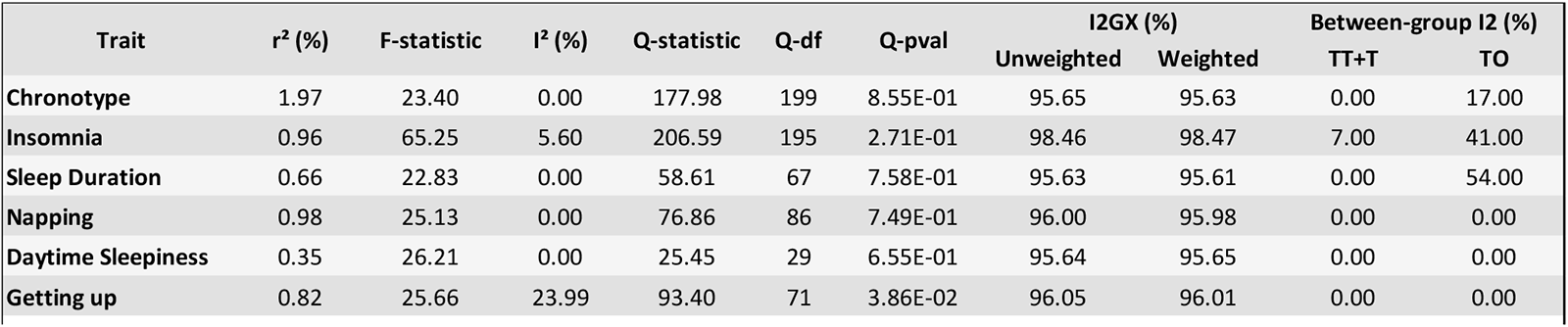
Summary of sensitivity results for sleep trait instruments. TT+T = tumour type and treatment; TO = treatment-only.

| Trait | r<sup>2</sup> (%) | F-statistic | I<sup>2</sup> (%) | Q-statistic | Q-df | Q-pval | I2GX (%) |  | Between-group I2 (%) |  |
| --- | --- | --- | --- | --- | --- | --- | --- | --- | --- | --- |
|  |  |  |  |  |  |  | Unweighted | Weighted | TT+T | TO |
| **Chronotype** | 1.97 | 23.40 | 0.00 | 177.98 | 199 | 8.55E-01 | 95.65 | 95.63 | 0.00 | 17.00 |
| **Insomnia** | 0.96 | 65.25 | 5.60 | 206.59 | 195 | 2.71E-01 | 98.46 | 98.47 | 7.00 | 41.00 |
| **Sleep Duration** | 0.66 | 22.83 | 0.00 | 58.61 | 67 | 7.58E-01 | 95.63 | 95.61 | 0.00 | 54.00 |
| **Napping** | 0.98 | 25.13 | 0.00 | 76.86 | 86 | 7.49E-01 | 96.00 | 95.98 | 0.00 | 0.00 |
| **Daytime Sleepiness** | 0.35 | 26.21 | 0.00 | 25.45 | 29 | 6.55E-01 | 95.64 | 95.65 | 0.00 | 0.00 |
| **Getting up** | 0.82 | 25.66 | 23.99 | 93.40 | 71 | 3.86E-02 | 96.05 | 96.01 | 0.00 | 0.00 |

We found evidence for morning preference (OR= 0.88, 95% CI= 0.82, 0.95) and ease of getting up (OR= 0.71, 95% CI= 0.55, 0.91) genetic instruments, but not others, influencing breast cancer occurrence (Figure 3A). Effect estimates for these two traits after using the Slope-Hunter correction [52] were consistent with those in the main analyses (Figure 3B), suggesting results were largely unaffected by index event bias.

**Figure 3.**
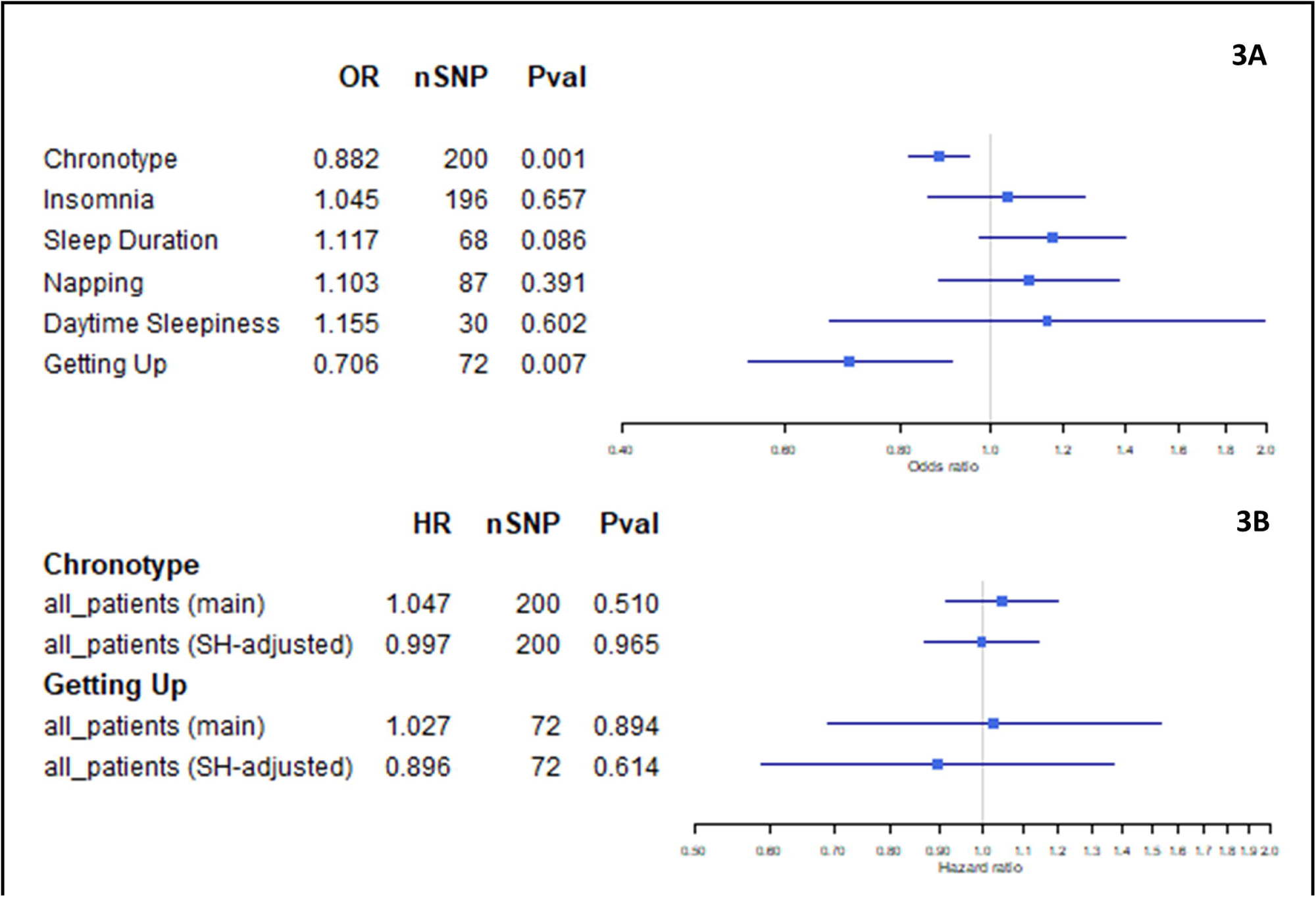
A) Two-sample MR results for sleep trait effect on breast cancer risk. B) Two-sample MR results for chronotype and ease of getting up effects on breast cancer mortality (unadjusted (main) vs slopehunter-adjusted (SH-adjusted)) at 15-years.

### Supplementary Analysis

We found some evidence that experiencing insomnia symptoms (compared to no symptoms) was protective against breast cancer specific mortality (i.e. prolonged survival) in patients with ER+/PR+ HER2-tumours who received no chemotherapy (HR= 0.18, 95% CI= 0.05, 0.68), as well as among patients given aromatase inhibitors (HR= 0.23, 95% CI= 0.07, 0.78), whilst this was not the case for those receiving other chemotherapies (**Supplementary figure 1**). There were no clear subgroup effects observed for the other sleep traits (**Supplementary figures 2-6**), although the highest between-group heterogeneity was found for the effect of sleep duration on breast cancer mortality when stratified by treatment-only (between-group I^2^=54%) (**Table 2**). However, once stratified, the number of deaths for each subgroup is low (n=314 to 2,298) (**Table 1**) and are therefore results are likely underpowered and must be interpreted with caution.

Following meta-analysis of subgroups, the highest between-group heterogeneity was found for the effect of sleep duration on breast cancer mortality when stratified by treatment-only (between-group I^2^=54%) (**Table 2**).

## DISCUSSION

### Summary of main findings

In this study, two-sample MR analyses were used to evaluate the causal role of sleep traits on breast cancer mortality. Initially we found some evidence linking daytime sleepiness with lower odds of overall breast cancer mortality, and insomnia symptoms with lower odds of mortality in women with ER+/PR+/HER2-tumours (receiving no chemotherapy) and all patients receiving aromatase inhibitors. Although our sensitivity analyses do not provide statistical evidence for a lack of robustness in our finding of a protective effect of daytime sleepiness on breast cancer mortality, given the behavioural links between sleep traits (i.e. people who have symptoms of insomnia, or shorter sleep duration, are likely to experience worse daytime sleepiness), it’s unlikely that we would see such a pronounced effect without seeing an effect from these other traits. Therefore, this result is likely due to type 1 error since it did not withstand Bonferroni correction. Overall, we were unable to demonstrate robust causal relationships between any of the sleep traits and either overall or subtype-specific breast cancer mortality.

These findings are important as sleep is vital to overall wellbeing and these findings provide some reassurance to patients with breast cancer, who often experience significant anxiety around their sleep [54], that there does not appear to be any harmful effects of sleep duration, quality or circadian timing on breast cancer-specific mortality.

### Previous studies

A recent study demonstrated that in both breast cancer patients and mouse models, generation and activity of circulating tumour cells is highly restricted to the ‘rest phase’ (i.e. sleep) of the circadian cycle, and that these cells have high proliferation rates and a greater metastatic ability compared to circulating tumour cells produced during the’ active phase’ (i.e. awake) [22]. As well as suggesting the need for time-controlled approaches treatment of breast cancer, these results imply that longer rest-phase i.e. longer sleep duration, may negatively impact survival outcomes. We did not find evidence for a causal relationship between lifelong propensity towards a longer sleep duration and breast cancer mortality and, while a protective effect of daytime sleepiness on breast cancer mortality was observed (which may indicate a prolonged ‘active phase’), this finding was not robust.

In support of our findings regarding the lack of effect of sleep duration on mortality, a recent observational study of 3047 women found that neither long (≥9 hours/night) nor short (≤6 hours/night) sleep duration was associated with breast cancer specific survival, provided the sleep was consistent (i.e. sleeping ≤6 hours every night, versus occasionally sleeping ≤6 hours when usual sleep duration is ≥6 hours) [55]. We acknowledge this analysis did not evaluate the same effects we have estimated, and it would not be possible to undertake the same analyses using MR with currently available data.

As part of our analysis, we have repeated and extending earlier work exploring the effect of habitual sleep patterns on breast cancer incidence. Our results for the effect of morning preference chronotype (OR=0.88, 95% CI=0.82, 0.95) and sleep duration (OR=1.12, 95% CI=0.98, 1.40) on breast cancer incidence are consistent with previously reported findings (OR=0.88, 95% CI=0.82, 0.93 and OR=1.19, 95% CI=1.02, 1.39 respectively) [27]. Our results for the effect of insomnia symptoms on breast cancer incidence (OR=1.05, 95% CI=0.86, 1.27) are largely consistent with previously reported findings (OR=0.80, 95% CI= 0.49, 1.31) in that both are imprecisely estimated and confidence intervals cross the null [27].

Our study took steps to explore effects of lifelong propensity to a particular sleep pattern on breast cancer mortality in subgroups of women on different treatments and observed some evidence that women with (lifelong) insomnia symptoms and ER+/PR+ had increased survival if they received no chemotherapy or aromatase inhibitors, compared to women on other chemotherapy. However, these subgroup analyses were extremely imprecise, with some subgroup analyses not possible because of small samples with low number of deaths in the subgroup and we consider them unreliable.

### Strengths and limitations

The use of two-sample MR to investigate the causal effect of each of our sleep traits on breast cancer is a key strength of this study and has enabled us to evaluate the effects of sleep in a large sample of breast cancer patients who did not have sleep traits directly measured. The use of newly released highly stratified breast cancer mortality outcome data of 91,686 patients to conduct these analyses has also provided granularity required to generate novel and clinically relevant results. In addition, we have thoroughly appraised our results in accordance with the main MR assumptions.

A further strength is the use of a series of sensitivity analyses to explore MR assumptions, including the use of Slope-Hunter to explore index-event bias [56] . It was not possible to use Slope-Hunter for outcomes stratified by treatment and/or tumour type because the necessary GWAS for breast cancer risk stratified in this way are not available, though given that our main results were largely unchanged following Slope-Hunter analyses, we think it is unlikely that these supplementary analyses are substantially biased.

Despite using the largest available dataset for breast cancer mortality, a limitation of this study is that sample sizes for subgroups were small and comparatively low-powered. While we found these subgroups a useful starting point for exploratory analyses, further research is required to fully understand the extent of sleep effects on breast cancer mortality, when stratified by tumour type and/or treatment. For example, while we observed some evidence that women with insomnia symptoms and ER+/PR+ had increased survival if they received no chemotherapy or aromatase inhibitors, compared to women on other chemotherapy, these subgroup analyses were extremely imprecise and therefore we cannot infer the effects of treatment-induced sleep disruption on breast cancer survival. While we found these subgroups a useful starting point for exploratory analyses, further research is required to fully understand the extent of sleep effects on breast cancer mortality, when stratified by tumour type and/or treatment. For example, it would be prudent to explore whether interventions which improve sleep in patients undergoing endocrine therapy also improve overall quality of life and adherence to treatment, which in turn, may serve to reduce mortality rates.

Furthermore, the outcome data in this study defines mortality as a binary outcome at a given timepoint. As such, we have been unable to look at the potential longitudinal effects of sleep on survival over time, for example over the course of treatment.

Another limitation of this study is the that the genetic variants are likely proxying the effects of life-long sleep behaviours, and therefore it is assumed that the genetic effects on are consistent pre- and post-diagnosis/treatment. Furthermore, the outcome data in this study defines survival as a binary outcome at a given timepoint. As such, we have been unable to look at the potential longitudinal effects of sleep on survival over time, for example over the course of treatment.

In addition, the sleep traits explored in this study were generated from self-report data and, while there is some evidence for moderate correlation between self-report and actigraphy-derived measures [30–33,57], recent findings have suggested these may not be as correlated as first thought, particularly for insomnia symptoms [49]. For insomnia, diagnostic criteria were not used to define this trait [37] and as such we were unable to say with any certainty that the participants had insomnia disorder, which would have strengthened this study.

### Further work

Given recent studies suggesting that melatonin may help to prevent tumour spread and worsening mortality rates [58], it would be useful if future large human studies were able to measure circadian variation in melatonin levels and perform a GWAS of these changes (using novel methods that are currently being developed [59]). If genetic instruments for such changes could be identified, this would be a step towards enabling the exploration of sleep traits effects that vary with age or following a cancer diagnosis.

In addition, it would be of interest to explore behavioural mechanisms, for example – medication compliance, within an MR framework. Aromatase inhibitor discontinuation has previously been studied in relation to musculoskeletal symptoms [60], and although it was not feasible to explore sleep difficulties in relation to AI discontinuation in our current study, we believe it would be of value if this becomes possible in the future. The relationship between sleep and adherence is likely complex – for example, those genetically pre-disposed to poor sleep traits (such as those that experience insomnia symptoms) could potentially be more tolerant of medications that reduce their sleep quality and may be more compliant. Sleep also has many other health implications with breast cancer and, in a population that is increasingly being diagnosed younger and benefiting from increased survival, taking account of the effects of sleep on all aspects of both physical and psychological well-being is paramount. Lastly, if it were possible to obtain sufficiently large sample sizes, we could undertake factorial MR of the combined effects of genetic instruments for the drug targets of chemotherapies (e.g. aromatase inhibitors) and the different sleep traits on survival.

### Public health and clinical implications

In contrast to recent findings from conventional multivariable regression analyses that prolonged sleep may negatively impact breast cancer survival [22,23], we found no robust evidence for an effect of any sleep trait on breast cancer specific mortality. We therefore would not advocate for shorter sleep duration in a public health setting, especially given that poor or disrupted sleep has a well-documented detrimental effect on numerous other outcomes, including mental and cardiometabolic health [61,62].

## CONCLUSION

In this study, we were unable to demonstrate a robust relationship between genetically predicted sleep traits and breast cancer survival and suggest caution is needed in messaging regarding the relationship between sleep habits and breast cancer survival. However, we cannot make any clear inferences about breast cancer treatment-induced sleep disruption, which requires investigation through well-conducted clinical studies with large sample sizes.

## Data Availability

This research was conducted using the UK Biobank Resource under application number 16391.
Summary data for the morning-preference, sleep duration, napping, daytime-sleepiness and ease of getting up in the morning GWAS used in this study are available at:
https://sleep.hugeamp.org/dinspector.html?dataset=GWAS_UKBB_eu
Summary data for the insomnia symptoms GWAS used in this study are available at:
https://pubmed.ncbi.nlm.nih.gov/35835914/
Summary data for breast cancer subtype survival GWAS used in this study are available at: https://bcac.ccge.medschl.cam.ac.uk/bcacdata/oncoarray/oncoarray-and-combined-summary-result/gwas-summary-results-survival-2021/

## FUNDING

BH is funded by an Above & Beyond breast cancer legacy grant from University Hospitals Bristol NHS Foundation Trust (www.aboveandbeyond.org.uk). LF is core-funded by the University of Strathclyde. OM is funded by the Business and Local Government Data Research Centre (BLG DRC) (ES/S007156/1). BH, TR, RMM, DAL and RCR are supported by or affiliated with the Medical Research Council Integrative Epidemiology Unit at the University of Bristol which is supported by the Medical Research Council (MC_UU_00032/01, MC_UU_00032/04, and MC_UU_00032/05) (www.mrc.ukri.org) and the University of Bristol. RMM and DAL are also supported by the National Institute for Health Research (NIHR) Bristol Biomedical Research Centre, which is funded by the National Institute for Health Research (NIHR) (www.nihr.ac.uk) and is a partnership between University Hospitals Bristol and Weston NHS Foundation Trust and the University of Bristol. DAL and RMM are NIHR Senior Investigators (NF-0616-10102, NIHR202411). RMM and RCR are supported by a Cancer Research UK (C18281/A29019) programme grant (the Integrative Cancer Epidemiology Programme). TR is supported by an NIHR Development and Skills and Enhancement Award (DSE) (NIHR 302363). The funders had no role in study design, data collection and analysis, decision to publish, or preparation of the manuscript. Department of Health and Social Care disclaimer: The views expressed are those of the author(s) and not necessarily those of the NHS, the NIHR or the Department of Health and Social Care.

## DATA AVAILABILITY

This research was conducted using the UK Biobank Resource under application number 16391.

Summary data for the morning-preference, sleep duration, napping, daytime-sleepiness and ease of getting up in the morning GWAS used in this study are available at: https://sleep.hugeamp.org/dinspector.html?dataset=GWAS_UKBB_eu

Summary data for the insomnia symptoms GWAS used in this study are available at: https://pubmed.ncbi.nlm.nih.gov/35835914/

Summary data for breast cancer subtype survival GWAS’ used in this study are available at: https://bcac.ccge.medschl.cam.ac.uk/bcacdata/oncoarray/oncoarray-and-combined-summary-result/gwas-summary-results-survival-2021/

## Notes

### Competing Interest Statement

DAL has received support from Roche Diagnostics and Medtronic Ltd for research unrelated to that presented here. TR has received grants from Daiichi-Sankyo and Amgen to attend educational workshops. All other authors have no competing interests to declare. The funders had no role in the design of the study; the collection, analysis, and interpretation of the data; the writing of the manuscript; and the decision to submit the manuscript for publication. The authors alone are responsible for the views expressed in this article.

### Author Declarations

The study was approved by the UKB data access committee, and informed consent was obtained from all participants. All participating UKB, 23andMe and BCAC studies were approved by their appropriate ethics or institutional review board and all participants provided informed consent.

