## Supplementary figures: 1-6 for "The Impact of Sleep on Breast Cancer-Specific Mortality: A Mendelian Randomisation Study"

| | HR | nSNP | Pval |
|---|---|---|---|
| 15 year mortality (all patients) | 0.949 | 196 | 0.738 |
| **Tumour-type and treatment** | | | |
| ER+ tumours (endocrine therapy) | 0.831 | 196 | 0.548 |
| ER- tumours (chemotherapy) | 0.89 | 196 | 0.793 |
| ER+/PR+ HER2- tumours (total) | 0.743 | 196 | 0.358 |
| ER+/PR+ HER2- tumours (chemotherapy) | 1.106 | 196 | 0.836 |
| ER+/PR+ HER2- tumours (no chemotherapy) | 0.178 | 196 | 0.012 * |
| ER+/PR+ HER2+ tumours (all therapies) | 1.88 | 196 | 0.295 |
| ER-/PR- HER2+ tumours (all therapies) | 1.013 | 196 | 0.986 |
| ER-/PR- HER2- tumours (all therapies) | 0.707 | 196 | 0.496 |
| **Treatment-only** | | | |
| All patients given tamoxifen | 1.176 | 196 | 0.623 |
| All patients given aromatase inhibitors | 0.232 | 196 | 0.018 * |
| All patients given CMF-like chemotherapy | 0.487 | 196 | 0.290 |
| All patients given taxanes | 1.147 | 196 | 0.825 |
| All patients given anthracyclines | 1.132 | 196 | 0.741 |

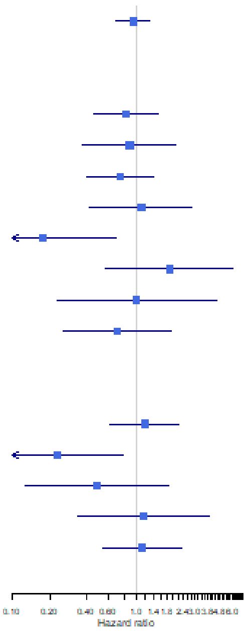


**Supplementary Figure 1.** Two-sample MR results for insomnia effect on breast cancer mortality overall and stratified by subgroups.
*Meets 0.05 Pval threshold
** Meets Bonferroni corrected Pval threshold of 0.0006

| | HR | nSNP | Pval |
|---|---|---|---|
| 15 year mortality (all patients) | 1.078 | 68 | 0.584 |
| **Tumour-type and treatment** | | | |
| ER+ tumours (endocrine therapy) | 1.279 | 68 | 0.328 |
| ER- tumours (chemotherapy) | 0.678 | 68 | 0.427 |
| ER+/PR+ HER2- tumours (total) | 1.01 | 68 | 0.968 |
| ER+/PR+ HER2- tumours (chemotherapy) | 0.696 | 68 | 0.367 |
| ER+/PR+ HER2- tumours (no chemotherapy) | 2.781 | 68 | 0.118 |
| ER+/PR+ HER2+ tumours (all therapies) | 1.163 | 68 | 0.812 |
| ER-/PR- HER2+ tumours (all therapies) | 0.866 | 68 | 0.833 |
| ER-/PR- HER2- tumours (all therapies) | 0.712 | 68 | 0.516 |
| **Treatment-only** | | | |
| All patients given tamoxifen | 1.263 | 68 | 0.420 |
| All patients given aromatase inhibitors | 2.674 | 68 | 0.071 |
| All patients given CMF-like chemotherapy | 0.28 | 68 | 0.061 |
| All patients given taxanes | 0.629 | 68 | 0.425 |
| All patients given anthracyclines | 0.756 | 68 | 0.399 |

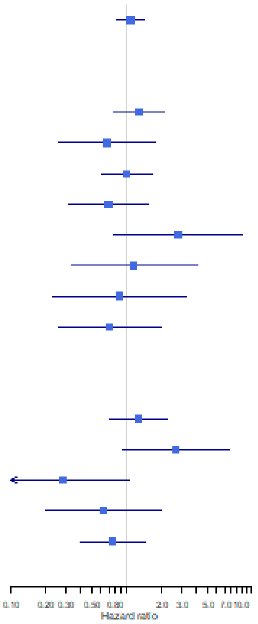


**Supplementary Figure 2.** Two-sample MR results for sleep duration effect on breast cancer mortality overall and stratified by subgroups.
*Meets 0.05 Pval threshold
** Meets Bonferroni corrected Pval threshold of 0.0006

| | HR | nSNP | Pval |
|---|---|---|---|
| 15 year mortality (all patients) | 0.336 | 30 | 0.013 * |
| **Tumour-type and treatment** | | | |
| ER+ tumours (endocrine therapy) | 0.419 | 30 | 0.372 |
| ER- tumours (chemotherapy) | 0.672 | 30 | 0.756 |
| ER+/PR+ HER2- tumours (total) | 0.331 | 30 | 0.182 |
| ER+/PR+ HER2- tumours (chemotherapy) | 1.742 | 30 | 0.679 |
| ER+/PR+ HER2- tumours (no chemotherapy) | 0.022 | 30 | 0.062 |
| ER+/PR+ HER2+ tumours (all therapies) | 0.544 | 30 | 0.745 |
| ER-/PR- HER2+ tumours (all therapies) | 0.019 | 30 | 0.068 |
| ER-/PR- HER2- tumours (all therapies) | 0.133 | 30 | 0.171 |
| **Treatment-only** | | | |
| All patients given tamoxifen | 0.648 | 30 | 0.725 |
| All patients given aromatase inhibitors | 0.772 | 30 | 0.882 |
| All patients given CMF-like chemotherapy | 0.476 | 30 | 0.748 |
| All patients given taxanes | 6.263 | 30 | 0.306 |
| All patients given anthracyclines | 1.234 | 30 | 0.848 |

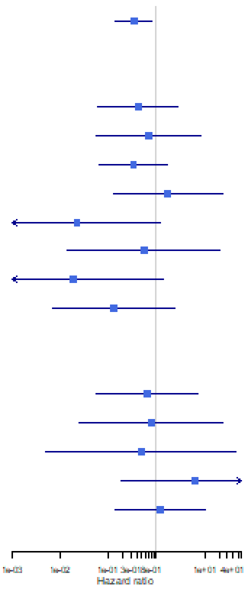


**Supplementary Figure 3.** Two-sample MR results for daytime sleepiness effect on breast cancer mortality overall and stratified by subgroups.

*Meets 0.05 Pval threshold

** Meets Bonferroni corrected Pval threshold of 0.0006

| | HR | nSNP | Pval |
|---|---|---|---|
| 15 year mortality (all patients) | 1.027 | 72 | 0.894 |
| **Tumour-type and treatment** | | | |
| ER+ tumours (endocrine therapy) | 2.072 | 72 | 0.066 |
| ER- tumours (chemotherapy) | 0.745 | 72 | 0.576 |
| ER+/PR+ HER2- tumours (total) | 1.572 | 72 | 0.176 |
| ER+/PR+ HER2- tumours (chemotherapy) | 1.838 | 72 | 0.248 |
| ER+/PR+ HER2- tumours (no chemotherapy) | 2.518 | 72 | 0.211 |
| ER+/PR+ HER2+ tumours (all therapies) | 0.684 | 72 | 0.585 |
| ER-/PR- HER2+ tumours (all therapies) | 1.75 | 72 | 0.525 |
| ER-/PR- HER2- tumours (all therapies) | 0.598 | 72 | 0.39 |
| **Treatment-only** | | | |
| All patients given tamoxifen | 0.838 | 72 | 0.699 |
| All patients given aromatase inhibitors | 0.909 | 72 | 0.903 |
| All patients given CMF-like chemotherapy | 0.893 | 72 | 0.888 |
| All patients given taxanes | 1.521 | 72 | 0.559 |
| All patients given anthracyclines | 1.447 | 72 | 0.458 |

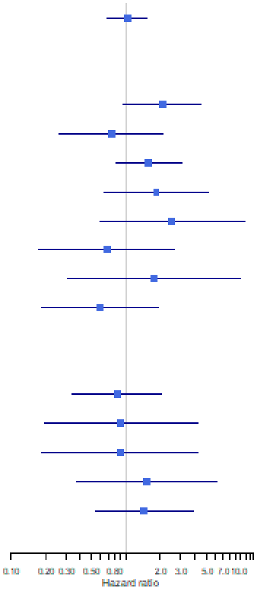


**Supplementary Figure 4.** Two-sample MR results for ease of getting up effect on breast cancer mortality overall and stratified by subgroups.

*Meets 0.05 Pval threshold

** Meets Bonferroni corrected Pval threshold of 0.0006

| | HR | nSNP | Pval |
|---|---|---|---|
| 15 year mortality (all patients) | 1.047 | 200 | 0.51 |
| **Tumour-type and treatment** | | | |
| ER+ tumours (endocrine therapy) | 0.985 | 200 | 0.903 |
| ER- tumours (chemotherapy) | 1.26 | 200 | 0.251 |
| ER+/PR+ HER2- tumours (total) | 1.08 | 200 | 0.553 |
| ER+/PR+ HER2- tumours (chemotherapy) | 0.956 | 200 | 0.824 |
| ER+/PR+ HER2- tumours (no chemotherapy) | 1.409 | 200 | 0.238 |
| ER+/PR+ HER2+ tumours (all therapies) | 0.743 | 200 | 0.26 |
| ER-/PR- HER2+ tumours (all therapies) | 1.308 | 200 | 0.433 |
| ER-/PR- HER2- tumours (all therapies) | 1.139 | 200 | 0.571 |
| **Treatment-only** | | | |
| All patients given tamoxifen | 0.836 | 200 | 0.215 |
| All patients given aromatase inhibitors | 1.169 | 200 | 0.583 |
| All patients given CMF-like chemotherapy | 1.248 | 200 | 0.488 |
| All patients given taxanes | 0.909 | 200 | 0.735 |
| All patients given anthracyclines | 1.32 | 200 | 0.097 |

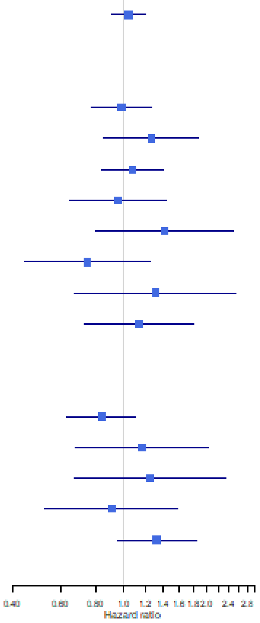


**Supplementary Figure 5.** Two-sample MR results for morning-preference chronotype effect on breast cancer mortality overall and stratified by subgroups.

*Meets 0.05 Pval threshold

** Meets Bonferroni corrected Pval threshold of 0.0006

| | HR | nSNP | Pval |
|---|---|---|---|
| 15 year mortality (all patients) | 0.942 | 87 | 0.794 |
| **Tumour-type and treatment** | | | |
| ER+ tumours (endocrine therapy) | 0.716 | 87 | 0.429 |
| ER- tumours (chemotherapy) | 0.814 | 87 | 0.758 |
| ER+/PR+ HER2- tumours (total) | 0.704 | 87 | 0.419 |
| ER+/PR+ HER2- tumours (chemotherapy) | 0.930 | 87 | 0.915 |
| ER+/PR+ HER2- tumours (no chemotherapy) | 0.402 | 87 | 0.377 |
| ER+/PR+ HER2+ tumours (all therapies) | 0.630 | 87 | 0.635 |
| ER-/PR- HER2+ tumours (all therapies) | 0.529 | 87 | 0.578 |
| ER-/PR- HER2- tumours (all therapies) | 0.407 | 87 | 0.330 |
| **Treatment-only** | | | |
| All patients given tamoxifen | 1.001 | 87 | 0.999 |
| All patients given aromatase inhibitors | 1.764 | 87 | 0.536 |
| All patients given CMF-like chemotherapy | 0.725 | 87 | 0.754 |
| All patients given taxanes | 0.800 | 87 | 0.812 |
| All patients given anthracyclines | 0.988 | 87 | 0.982 |

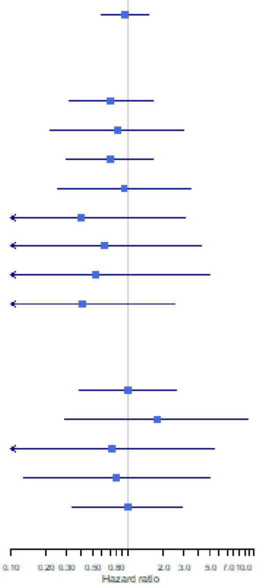


**Supplementary Figure 6.** Two-sample MR results for daytime napping effect on breast cancer mortality overall and stratified by subgroups.

*Meets 0.05 Pval threshold

** Meets Bonferroni corrected Pval threshold of 0.0006
