## Supplementary methods: A-D for "The Impact of Sleep on Breast Cancer-Specific Mortality: A Mendelian Randomisation Study"

### *Supplementary Methods A: Sleep Trait Questionnaires in UK Biobank*

At baseline, participants completed a touchscreen questionnaire, which included questions about their sleep behaviours.

Chronotype (morning or evening preference) was assessed in the question “Do you consider yourself to be?” with one of six possible answers: “Definitely a ‘morning’ person,” “More a ‘morning’ than ‘evening’ person,” “More an ‘evening’ than a ‘morning’ person,” “Definitely an ‘evening’ person,” “Do not know,” or “Prefer not to answer”, from which we derived a five category variable for chronotype where “Definitely a ‘morning’ person” and “Definitely an ‘evening’ person” define either extreme.

Sleep duration was assessed by asking: “About how many hours sleep do you get in every 24 hours? (Including naps).” The answer could only contain integer values.

To assess insomnia symptoms, participants were asked: “Do you have trouble falling asleep at night or do you wake up in the middle of the night?” where responses “Never/rarely”, “Sometimes” or “Usually” were coded as a three-category variable.

Frequency of napping was assessed based on the question: “Do you have a nap during the day?”, with possible responses “Never/rarely”, “Sometimes” and “Usually” coded as a three-category variable.

The frequency of daytime-sleepiness using the question: “How likely are you to doze off or fall asleep during the daytime when you don’t mean to? (e.g.: when working, reading, or driving)”, with the answers coded as a four-category variable “never”, “sometimes”, “often”, or “all of the time”.

Ease of getting up in the morning was assessed using the question: "On an average day, how easy do you find getting up in the morning?", with the answers coded as a four-category variable “not at all easy”, “not very easy”, “fairly easy”, or “very easy”.

For all UKB-derived variables, answers “Prefer not to answer” were coded as missing.

*Supplementary Methods B: Genotyping and Imputation (UKB)*

Individuals were genotyped using the UK BiLEVE array (N=49,979) and the UK Biobank axiom array (N=438,398), with the full data release comprising of N=488,377 successfully genotyped samples. Multiallelic SNPs or those with a MAF of ≤1% were removed prior to phasing, which was then performed using a modified version of SHAPEIT2 [2]. Genotype imputation was performed using IMPUTE2 algorithms [3,4]. Further details for pre-imputation quality control, phasing and imputation are described elsewhere [5].

Individuals with mismatch between genetic sex and reported sex or with sex-chromosome aneuploidy were excluded from the analysis (N=814). Remaining samples were restricted to individuals of 'European' ancestry, defined using four principal components provided by UKB (N=464,708) [6].

GWAS in this study were conducted using the linear mixed model (LMM) association method as implemented in BOLT-LMM (v2.3) [7]. Population structure was modelled using directly genotyped SNPs, obtained after filtering on MAF > 0.01; Hardy-Weinberg equilibrium p-value < 0.0001 and LD pruning to an r2 threshold of 0.001 using PLINKv2.00.

*Supplementary Methods C: Sleep Trait Questionnaires in 23andMe*

23andMe is a personal genetics company[1], in which participants are given the option to consent to their data being used in research. In 23andme, an insomnia phenotype was determined through completion of questionnaires through online surveys.

Participants were asked to answer one or more questions related to seven sleep-related traits.

Participants with positive response to any of the following questions were considered as cases: 1) 'Have you ever been diagnosed with, or treated for, insomnia?', 2) 'Were you diagnosed with insomnia?', 3) 'Have you ever been diagnosed by a doctor with any of the following neurological conditions?' (Sleep disturbance), 4) 'Do you routinely have trouble getting to sleep at night?', 5) 'What sleep disorders have you been diagnosed with? Please select all that apply.' (Insomnia, trouble falling or staying asleep), 6) 'Have you ever taken these medications?' (Prescription sleep aids) and 7) 'In the last 2 years, have you taken any of these medications?' (Prescription sleep aids). Participants who did not provide either positive or uncertain answer ('I don't know' or 'I am not sure') in any of the 7 questions nor the followings are considered as controls: 1) 'Have you ever been diagnosed with, or treated for, any of the following conditions?' (Insomnia; Narcolepsy; Sleep apnoea; Restless leg syndrome), 2) 'In the past 12 months, have you been newly diagnosed with any of the following conditions by a medical professional?' (Insomnia; Sleep apnoea; Migraines), 3) 'Have you ever been diagnosed with or treated for any of the following conditions?' (Posttraumatic stress disorder; Autism; Asperger's; Sleep disorder), 4) 'Have you ever been diagnosed with or treated for a sleep disorder?', and 5) 'Have you ever been diagnosed with or treated for any of the following conditions?' (A sleep disorder).

#### *Supplementary Methods D: Genotyping and Imputation (BCAC)*

Patients were genotyped with iCOGS and OncoArray arrays [8,9], and were then restricted to variants with a minor allele frequency (MAF) > 0.01 and to participants of 'European' ancestry, based on genotype data. Non-genotyped variants were either imputed based on the 1000 Genomes Project Phase 3 (October 2014) release as reference panel, or using a reference panel from the Haplotype Reference Consortium (HRC) [10], to improve imputation quality for rarer variants.

Further details of methods related to genotyping and genotype imputation have been described previously [11–13].
